# SARS-CoV-2 and endemic coronaviruses: Comparing symptom presentation and severity of symptomatic illness among Nicaraguan children

**DOI:** 10.1101/2021.12.09.21267537

**Authors:** Aaron M Frutos, John Kubale, Guillermina Kuan, Sergio Ojeda, Nivea Vydiswaran, Nery Sanchez, Miguel Plazaola, May Patel, Roger Lopez, Angel Balmaseda, Aubree Gordon

**Author notes:** Corresponding author (AG). These authors contributed equally (Angel Balmaseda, Aubree Gordon).

## Abstract

It has been proposed that as SARS-CoV-2 transitions to endemicity, children will represent the greatest proportion of SARS-Co-V-2 infections as they currently do with endemic coronavirus infections. While SARS-CoV-2 infection severity is low for children, it is unclear if SARS-CoV-2 infections are distinct in symptom presentation, duration, and severity from endemic coronavirus infections in children. We compared symptom risk and duration of endemic human coronavirus (HCoV) infections from 2011-2016 with SARS-CoV-2 infections from March 2020-September 2021 in a Nicaraguan pediatric cohort. Blood samples were collected from study participants annually in February-April. Respiratory samples were collected from participants that met testing criteria. Blood samples collected in were tested for SARS-CoV-2 antibodies and a subset of 2011-2016 blood samples from four-year-old children were tested for endemic HCoV antibodies. Respiratory samples were tested for each of the endemic HCoVs from 2011-2016 and for SARS-CoV-2 from 2020-2021 via rt-PCR. By April 2021, 854 (49%) cohort participants were ELISA positive for SARS-CoV-2 antibodies. Most participants had antibodies against one alpha and one beta coronavirus by age four. We observed 595 symptomatic endemic HCoV infections from 2011-2016 and 121 symptomatic with SARS-CoV-2 infections from March 2020-September 2021. Symptom presentation of SARS-CoV-2 infection and endemic coronavirus infections were very similar, and SARS-CoV-2 symptomatic infections were as or less severe on average than endemic HCoV infections. This suggests that, for children, SARS-CoV-2 may be just another endemic coronavirus. However, questions about the impact of variants and the long-term effects of SARS-CoV-2 remain.

## Introduction

As severe acute respiratory syndrome coronavirus 2 (SARS-CoV-2) transitions to global endemicity, there are many questions about how that will occur. Over time, children will represent the greatest proportion of primary SARS-CoV-2 infections as adults gain immunity from natural infection or vaccination. (1) It is well established that pediatric risk of severe illness and death is much lower than that for adults. (2, 3) Differences in immune response between adults and children likely provide children better protection against severe SARS-CoV-2 infections. (3–5) Previous research found no differences in severity between SARS-CoV-2 infections and influenza A and B among children. (6, 7) However, it is unknown if, in children, SARS-CoV-2 infections are distinct in disease presentation and severity from endemic human coronavirus (HCoV) infections.

As of January 2022, only the Pfizer vaccine (for those age 5 or older) and the Moderna vaccine (for those 12 or older) are recommended by the World Health Organization’s Strategic Advisory Group of Experts (SAGE) for use in children and adolescents; multiple other vaccines, including the Cuban Soberana 02 and Abdala, have been approved for use in children by individual countries, including in Nicaragua.(8–10) If SARS-COV-2 infections are more severe, routine pediatric vaccination will be necessary to reduce excess mortality and morbidity. If not, vaccine-induced immunity should prevent severe disease while allowing for transmission to facilitate frequent immune boosting. (1) To determine if SARS-CoV-2 infections have distinct disease presentation from endemic HCoV infections in children, we compare symptomatic SARS-CoV-2 and endemic HCoV infection symptomology and severity in a prospective, community-based pediatric cohort in Managua, Nicaragua from 2011-2016 and 2020-2021

## Methods

Institutional review boards at the Nicaraguan Ministry of Health and the University of Michigan approved this study. Parents/guardians of participants provided written informed consent and participants aged ≥6 years provided verbal assent. The Nicaraguan Pediatric Influenza Cohort Study (NPICS) is a prospective cohort study that began in 2011 and continues today. Children ages 0-14 years who live in District 2 of Managua, Nicaragua and within the catchment area of Health Center Sócrates Flores Vivas were eligible to participate. Participants live in a tropical, urban environment and are representative of children in the larger Managua area. Participants included in this analysis were members of NPICS between 2011-2016 or March 2020 to September 2021. School-aged children attended school throughout the study period.

Study staff collected blood samples from each study participant between February and April each year. To confirm our assumption that endemic HCoV infection rates are high in the cohort, we tested a random subset of 100 blood samples from four-year-old’s from 2011-2016; using protocols for an enzyme-linked immunosorbent assay (ELISA) developed at Mount Sinai (11), we tested samples for IgG antibody response to the spike protein for each of the four endemic HCoVs (alpha: NL63, and 229E, beta: OC43 and HKU1). To confirm that SARS-CoV-2 infections rates were also high, we tested the 2020 and 2021 blood samples in pairs for SARS-CoV-2 IgG antibodies as described previously. If a 2020 annual sample was positive for SARS-CoV-2, the child’s 2019 annual sample was also run. Children that were positive in 2019 were not considered SARS-CoV-2 positive. (12)

Parents/guardians agreed to bring participants to the health center at the first signs of fever. Study personnel collected a respiratory swab from participants if they met the testing criteria: feverishness for participants under two years old; measured fever/feverishness and cough, sore throat, or rhinorrhea; severe respiratory symptoms such as apnea or chest indrawing; hospitalization with respiratory symptoms or sepsis. NPICS testing criteria expanded in June 2020 to capture mild symptomatic SARS-CoV-2 cases; however, this analysis is limited to those meeting the original testing criteria to ensure comparability. We used real-time reverse transcription polymerase-chain reaction (RT-PCR) to test respiratory samples from 2011-2016 for each of the four endemic HCoV, influenza A and B, respiratory syncytial virus (RSV, subtypes A and B), and human metapneumovirus (HMpV) and samples from March 2020-September 2021 for SARS-CoV-2 and influenza. (13, 14)

Study clinicians record all participant symptoms, prescriptions, and diagnoses during each clinic visit and subsequent medical appointments using standardized forms. Parents/guardians report participant symptoms for each day since illness onset to study clinicians at the initial and subsequent clinic visits until symptom resolution. We considered symptoms to be associated with an infection if they occurred within 28 days of symptom onset. Study parents/guardians reported on the following symptoms 2011-2016 and 2020-2021: feverishness, cough, rhinorrhea, nasal congestion, loss of appetite, myalgia, arthralgia, and rapid breathing. We considered participants who presented to the clinic with rapid breathing, rhonchi, indrawing, wheezing, or shortness of breath as having abnormal breathing. We defined acute lower respiratory infections (ALRI) as physician diagnosed cases of pneumonia, bronchiolitis, bronchitis, or bronchial hyperreactivity or elevated respiratory rate based on age: ≥ 60 breaths/minute for < 2 months, ≥ 50 breaths/minute for 2-11 months, ≥ 40 breaths/minute for 12-59 months, ≥ 25 breaths/minute for ≥ 60 months (15). We also evaluated whether participants with ALRI were prescribed antibiotics (amoxicillin, penicillin, other) within 28 days of infection. Data collection forms for the above signs and symptoms were consistent between 2011-2016 and 2020-2021.

To compare risk of symptoms, we calculated symptom specific risk differences between SARS-CoV-2 infections and endemic HCoV infections, overall and stratified by the following age groups: 0-4, 5-9, and 10-14 years. We also stratified the results by sex and endemic HCoV species. Using upset plots, we explored which signs/symptoms tended to present together. We also assessed symptom duration by comparing the time between each specific symptom onset and the last day participants presented with that symptom. We plotted symptom duration using boxplots and compared the distribution of symptom duration between SARS-CoV-2 and endemic HCoVs using the Mann Whitney U test.

We used SAS version 9.4 (SAS Institute Inc.) to calculate risk differences and ratios and R version 4.1.0 to create figures and conduct all other analyses.

## Results

There were 3,220 participants active in NPICS during the included years: 2,576 from 2011-2016 and 1,942 from March 2020-September 2021. On average, there were 1,792 active participants per year. Our assumption was that detected symptomatic infections consist of only a small proportion of total SARS-CoV-2 and endemic HCoV infections that occurred in the cohort children. Specifically, of the 1,942 children in the cohort from March 2020-September 2021, 1,455 in 2020 and 1,743 in 2021 had a blood sample collected that was tested via ELISA for SARS-CoV-2 antibodies. Only 23 (1.6%) tested positive for SARS-CoV-2 antibodies in 2020 while 854 (49%) tested positive in 2021 indicating that SARS-CoV-2 infections were very high in the cohort. We found that 94% of our randomly selected subset of four-year-olds from 2011-2016 had an antibody response to at least one alpha and one beta HCoV, confirming our assumption of high endemic HCoV infection rates in the community. Antibody response prevalence was highest for OC43 (99%) followed by HKU1 (86%), NL63 (83%), and then 229E (74%) (Table 1).

**Table 1:** Endemic HCoVs ELISA Results, % Positive.

|  | **Total (n=100)** |
| --- | --- |
| **Alpha** | 94% |
| NL63 | 83% |
| 229E | 74% |
| **Beta** | 100% |
| OC43 | 99% |
| HKU1 | 86% |

Within this cohort we observed high infection rates of both endemic HCoVs and SARS-CoV-2. That there were 595 RT-PCR+ symptomatic endemic HCoV infections from 2011-2016 and 121 RT-PCR+ symptomatic SARS-CoV-2 infections from March 2020-September 2021 again suggests that symptomatic cases represent only a small proportion of overall infections and likely, the more severe infections. Most endemic HCoV, 432 (73%), and SARS-CoV-2, 59 (49%), infections occurred in participants <5 years. Fever, cough, rhinorrhea, and congestion were the most common symptoms for both endemic HCoVs and SARS-CoV-2 symptomatic infections. Cough, rhinorrhea, and abnormal breathing was more common among endemic HCoV infections while measured fever and headache was more common among SARS-CoV-2 infections. Among SARS-CoV-2 symptomatic infections, 4 (3%) were acute lower respiratory infections compared to 98 (16%) endemic HCoV cases (p=<0.0001; Table 2). Among those with ALRI, there was not a difference in antibiotic prescription between SARS-CoV-2 and endemic HCoV infections.

**Table 2:** Study Participants and Symptom Prevalence.

|  | Endemic HCoVs<br>(n=595) | SARS-CoV-2<br>(n=121) | p-value* |
| --- | --- | --- | --- |
| **Age Group (%)** |  |  | <.0001 |
| 0-4 | 432 (73) | 59 (49) |  |
| 5-9 | 110 (18) | 29 (24) |  |
| 10-14 | 53 (9) | 33 (27) |  |
| **Symptoms (%)** |  |  |  |
| Measured fever | 272 (46) | 68 (56) | 0.035 |
| Cough | 524 (88) | 85 (70) | <0.001 |
| Rhinorrhea | 505 (85) | 93 (77) | 0.030 |
| Congestion | 276 (46) | 45 (37) | 0.064 |
| Sore throat | 145 (24) | 36 (30) | 0.221 |
| Headache | 81 (14) | 38 (31) | <0.001 |
| Loss of appetite | 142 (24) | 31 (26) | 0.681 |
| Diarrhea | 64 (11) | 14 (12) | 0.793 |
| Hospitalized | 23 (4) | 6 (5) | 0.578 |
| Abnormal breathing | 108 (18) | 9 (7) | 0.004 |
| Acute lower respiratory infection | 107 (18) | 7 (6) | <0.001 |
| ALRI and prescribed antibiotics† | 79 (74) | 6 (85) | 0.484 |

\* p-value from chi-square test
† % represents % of ALRI cases

Because the age distribution varied between endemic HCoVs and SARS-CoV-2, with a greater proportion of infections occurring in older children with SARS-CoV-2, we examined signs and symptoms by age group. Across age groups, participants with symptomatic endemic HCoV infections displayed greater risk of cough compared to those with symptomatic SARS-CoV-2 infections. Among participants aged <5 years, symptomatic endemic HCoV infections showed greater risk of rhinorrhea, congestion, abnormal breathing, and ALRI than symptomatic SARS-CoV-2 infections, while SARS-CoV-2 exhibited greater risk of measured fever (Fig 1, Table 3). Notably, among those under 5, symptomatic endemic HCoV infection was associated with greater risk of ALRI even after excluding participants that also tested positive for influenza A, influenza B, RSV, or HMpV. We observed no difference in risk hospitalization between SARS-CoV-2 and endemic HCoV symptomatic infections in each age group. These results were consistent after stratifying by sex (S1-S2 Fig), or endemic HCoV species (S3-S6 Fig).

**Table 3:** Symptom Risk between Endemic HCoV and SARS-CoV-2 by Age Group.

| **0-4** |  |  |  |  |
| --- | --- | --- | --- | --- |
|  | Endemic HCoVs (%) | SARS-CoV-2 (%) | Risk Difference (95% CI) | Risk Ratio (95% CI) |
| **Measured fever** | 218 (50) | 44 (75) | -0.24 (-0.36, -0.12) | 0.68 (0.57, 0.81) |
| **Cough** | 382 (88) | 42 (71) | 0.17 (0.05, 0.29) | 1.24 (1.05, 1.47) |
| **Rhinorrhea** | 378 (88) | 42 (71) | 0.16 (0.04, 0.28) | 1.23 (1.04, 1.45) |
| **Congestion** | 241 (50) | 21 (36) | 0.14 (0.09, 0.27) | 1.39 (0.97, 1.99) |
| **Sore throat** | 66 (15) | 6 (10) | 0.05 (-0.03, 0.14) | 1.50 (0.68, 3.31) |
| **Headache** | 23 (5) | 4 (7) | -0.01 (-0.08, 0.05) | 0.79 (0.28, 2.19) |
| **Loss of appetite** | 116 (27) | 19 (32) | -0.05 (-0.18, 0.07) | 0.83 (0.56, 1.25) |
| **Diarrhea** | 58 (13) | 9 (15) | -0.02 (-0.12, 0.08) | 0.88 (0.46, 1.68) |
| **Hospitalized** | 17 (4) | 4 (7) | -0.03 (-0.10, 0.04) | 0.58 (0.20, 1.67) |
| **Abnormal breathing** | 95 (22) | 4 (7) | 0.15 (0.08, 0.23) | 3.24 (1.24, 8.49) |
| **ALRI** | 89 (21) | 4 (7) | 0.14 (0.6, 0.21) | 3.04 (1.16, 7.97 ) |
| **ALRI\*** | 69 (19) | 4 (7) | 0.12 (0.04,0.19) | 3.74 (1.04, 7.22) |
| **5-9** |  |  |  |  |
|  | Endemic HCoVs (%) | SARS-CoV-2 (%) | Risk Difference (95% CI) | Risk Ratio (95% CI) |
| **Measured fever** | 39 (35) | 12 (41) | -0.06 (-0.26, 0.14) | 0.86 (0.52, 1.41) |
| **Cough** | 99 (90) | 21 (72) | 0.18 (0.00, 0.35) | 1.24 (0.98, 1.57) |
| **Rhinorrhea** | 87 (79) | 25 (86) | -0.07 (-0.22, 0.08) | 0.92 (0.77, 1.09) |
| **Congestion** | 42 (38) | 14 (48) | -0.10 (-0.10, 0.30) | 0.79 (0.51, 1.23) |
| **Sore throat** | 52 (47) | 12 (41) | 0.06 (-0.14, 0.26) | 1.14 (0.71, 1.84) |
| **Headache** | 35 (32) | 14 (48) | -0.16 (-0.37, 0.04) | 0.66 (0.41, 1.04) |
| **Loss of appetite** | 19 (17) | 7 (24) | -0.07 (-0.24, 0.10) | 0.72 (0.33, 1.54) |
| **Diarrhea** | 4 (4) | 3 (10) | -0.07 (-0.18, 0.05) | 0.35 (0.08, 1.48) |
| **Hospitalized** | 5 (5) | 2 (7) | -0.02 (-0.12, 0.08) | 0.66 (0.13, 3.23) |
| **Abnormal breathing** | 10 (9) | 2 (7) | 0.02 (-0.08, 0.12) | 1.32 (0.31, 5.69) |
| **ALRI** | 15 (14) | 3 (10) | 0.03 (-0.10, 0.16) | 1.32 (0.41, 4.25) |
| **ALRI\*** | 12 (13) | 3 (10) | 0.03 (-0.10, 0.16) | 1.26 (0.38, 4.16) |
| **10-14** |  |  |  |  |
|  | Endemic HCoVs (%) | SARS-CoV-2 (%) | Risk Difference (95% CI) | Risk Ratio (95% CI) |
| **Measured fever** | 15 (28) | 12 (36) | -0.08 (-0.28, 0.12) | 0.78 (0.42, 1.45) |
| **Cough** | 43 (81) | 22 (67) | 0.14 (-0.05, 0.34) | 1.22 (0.93, 1.60) |
| **Rhinorrhea** | 40 (75) | 26 (79) | -0.03 (-0.21, 0.15) | 0.96 (0.76, 1.21) |
| **Congestion** | 20 (38) | 10 (30) | 0.07 (-0.13, 0.28) | 1.25 (0.67, 2.32) |
| **Sore throat** | 27 (51) | 18 (55) | -0.04 (-0.25, 0.18) | 0.93 (0.62, 1.41) |
| **Headache** | 23 (43) | 20 (61) | -0.17 (-0.39, 0.04) | 0.25 (0.47, 1.08) |
| **Loss of appetite** | 7 (13) | 5 (15) | -0.02 (-0.17, 0.13) | 0.87 (0.30, 2.52) |
| **Diarrhea** | 2 (4) | 2 (6) | -0.02 (-0.12, 0.07) | 0.62 (0.09, 4.21) |
| **Hospitalized** | 1 (2) | 0 | 0.02 (-0.02, 0.06) | - |
| **Abnormal breathing** | 3 (6) | 3 (9) | -0.03 (-0.15, 0.08) | 0.62 (0.13, 2.90) |
| **ALRI** | 3 (4) | 0 | 0.06 (-0.01, 0.12) | - |
| **ALRI\*** | 3 (6) | 0 | 0.06 (-0.01, 0.13) | - |

\*Excluding influenza A, influenza B, RSV, and HMPV coinfections

**Fig 1:**
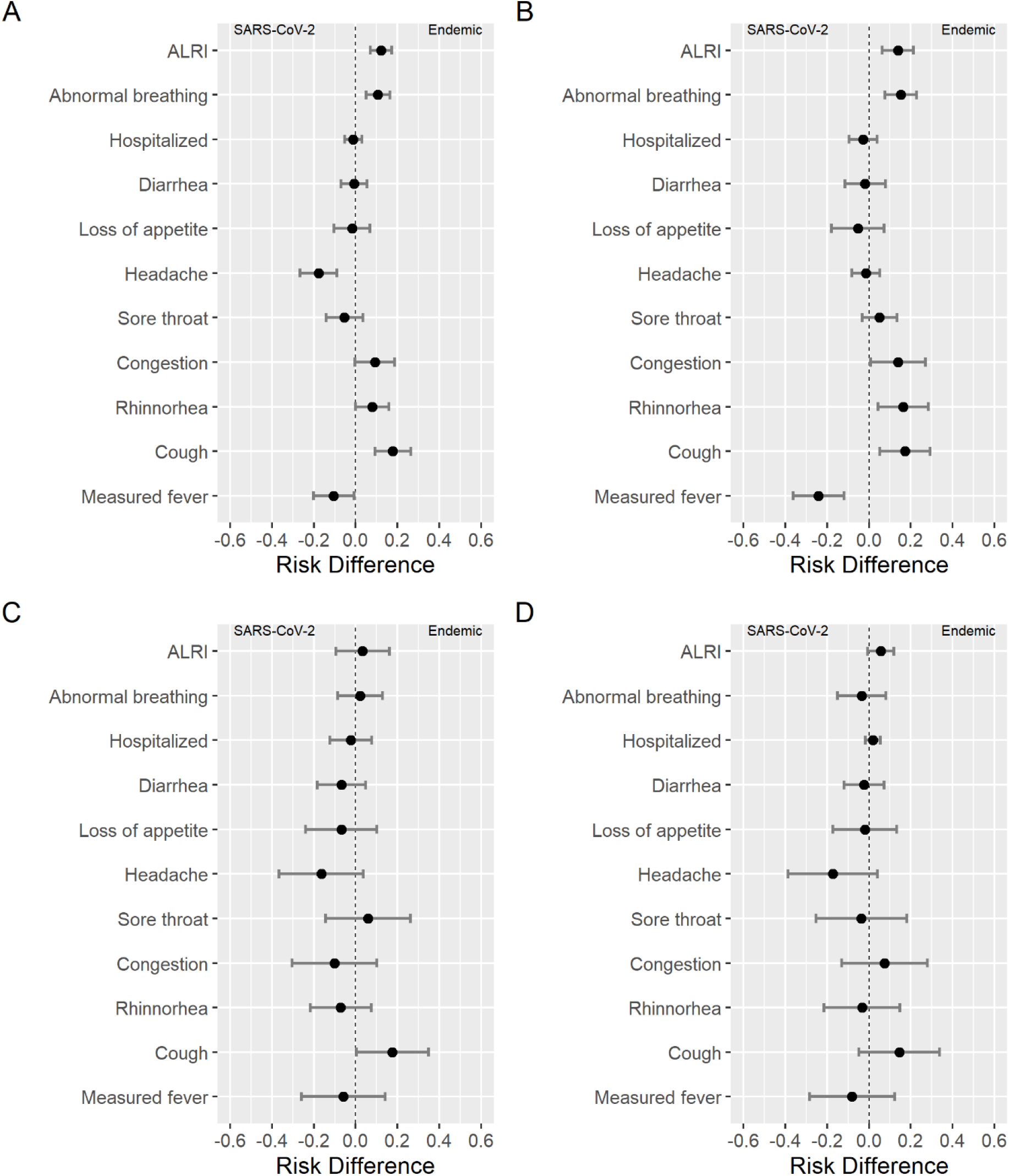
Symptom Risk Difference between Endemic HCoVs and SARS-CoV-2. A: All participants. B: Ages 0-4 years. C: Ages 5-9 years. D Ages: 10-14 years.

For both endemic HCoVs and SARS-CoV-2, we found that cough, rhinorrhea, and sore throat frequently presented together. Loss of appetite appeared in common symptom groupings for endemic HCoVs, while headache was part of more common groupings for SARS-CoV-2 (Fig 2). We did find that among participants aged 0-4 years loss of appetite lasted longer and among participants aged 5-9 and 10-14, cough lasted longer for SARS-CoV-2 infections., (Fig 3, Table 4).

**Table 4:**
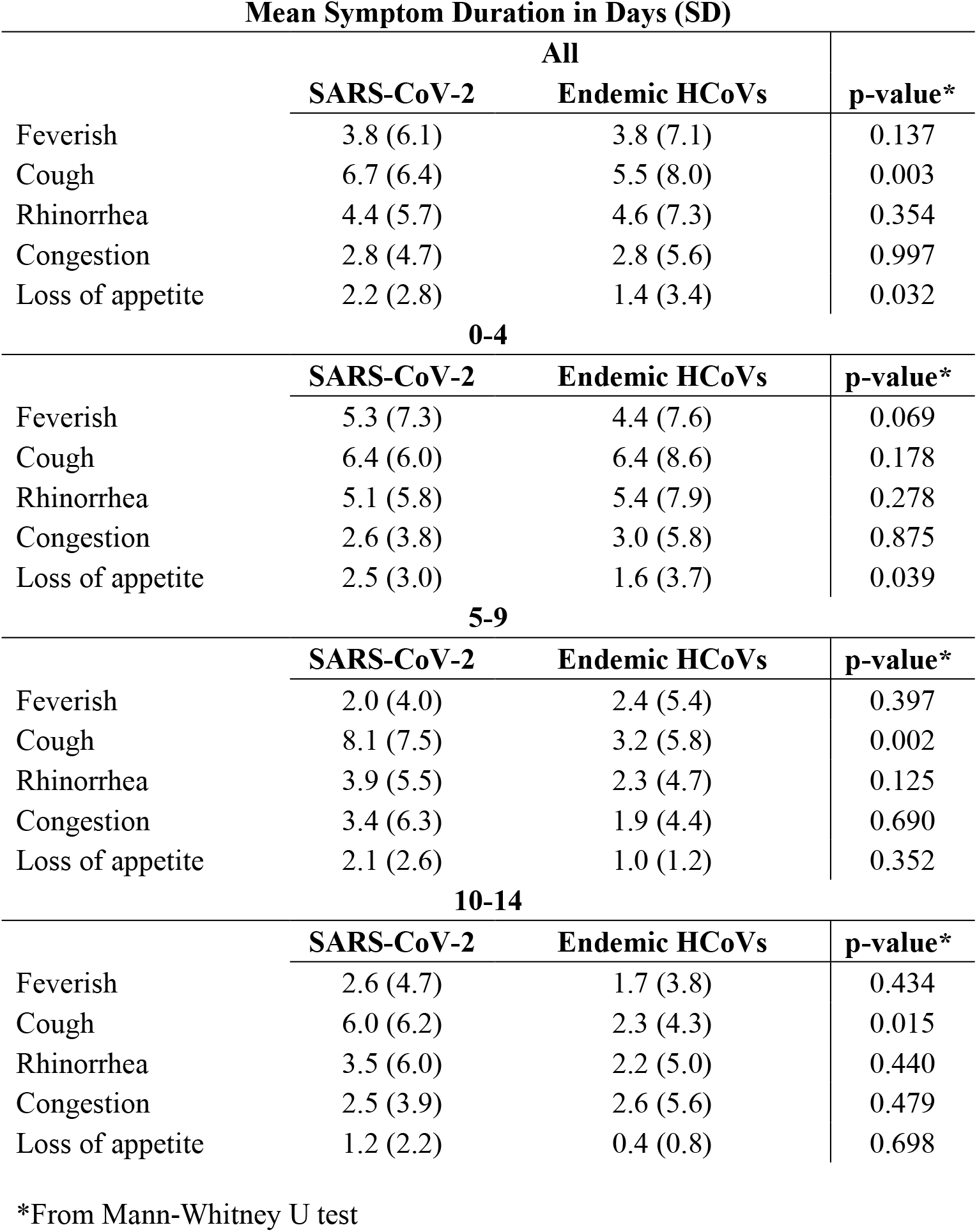
Symptom Duration Comparison: SARS-CoV-2, Endemic HCoVs.

**Fig 2.**
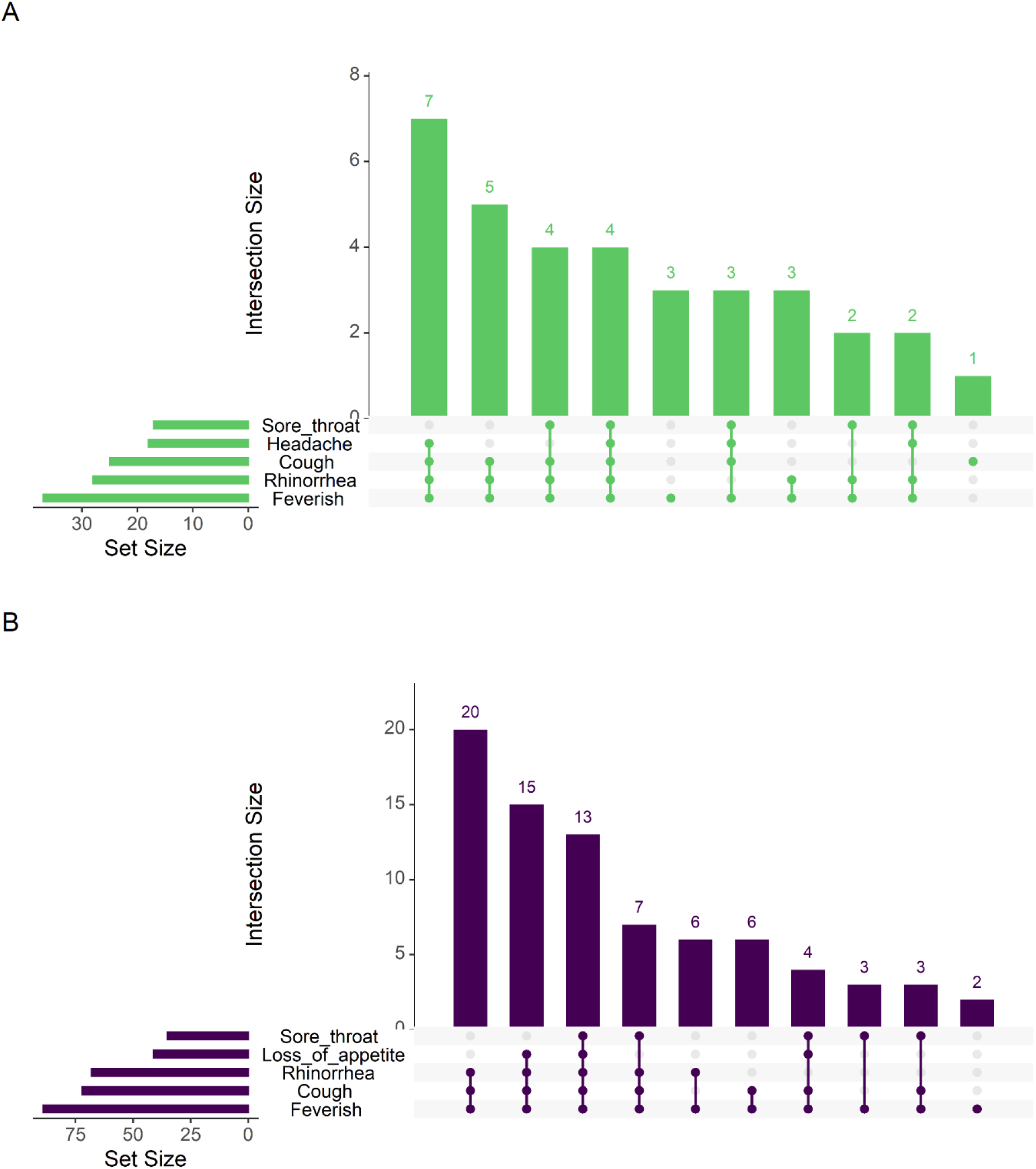
Comparison of Common Symptom Groupings between Symptomatic Endemic HCoVs and SARS-CoV-2 Infections. A: Upset plot of symptom groupings for symptomatic SARS-CoV-2 infections. B. Upset plot of symptom groupings for symptomatic endemic HCoV infections.

**Fig 3.**
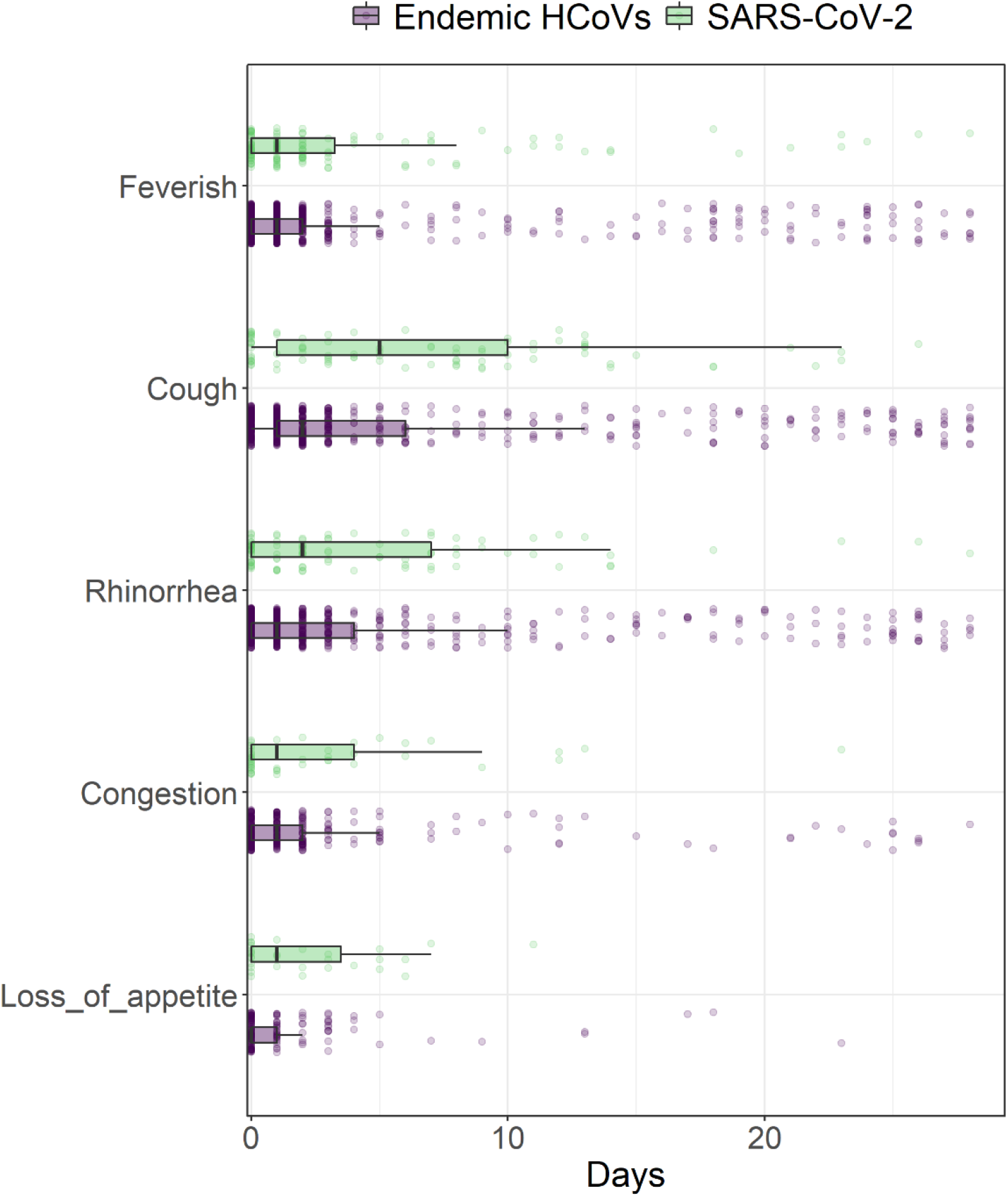
Comparison of Symptom Duration between Symptomatic Endemic HCoVs and SARS-CoV-2 Infections.

To assess the potential impact of variants on symptoms, we compared symptoms for SARS-CoV-2 infections from 2020 prior to the global emergence of variants and 2021 when delta, gamma, and lambda strains circulated in the cohort area and found no difference in presentation by year. Feverishness, rhinorrhea, cough, headache, and sore throat where the most common symptoms for SARS-CoV-2 cases in both 2020 and 2021. (Fig 4). All observed cases of ALRI associated with SARS-CoV-2 occurred in 2020. However, we did find that rhinorrhea lasted longer in SARS-CoV-2 cases from 2021 compared to 2020 (Table 5).

**Table 5:** Symptom Duration Comparison: SARS-CoV-2 by Year.

|  | Mean Symptom Duration<br>in Days (SD) |  | p-value* |
| --- | --- | --- | --- |
|  | 2020 | 2021 |  |
| Feverish | 4.1 (7.4) | 3.6 (5.0) | 0.129 |
| Cough | 6.8 (8.0) | 6.7 (5.5) | 0.382 |
| Rhinorrhea | 2.7 (5.0) | 5.4 (6.0) | 0.004 |
| Congestion | 1.6 (2.3) | 3.3 (5.2) | 0.378 |
| Loss of appetite | 3.5 (3.8) | 1.6 (1.9) | 0.172 |

\*From Mann-Whitney U test

**Fig 4.**
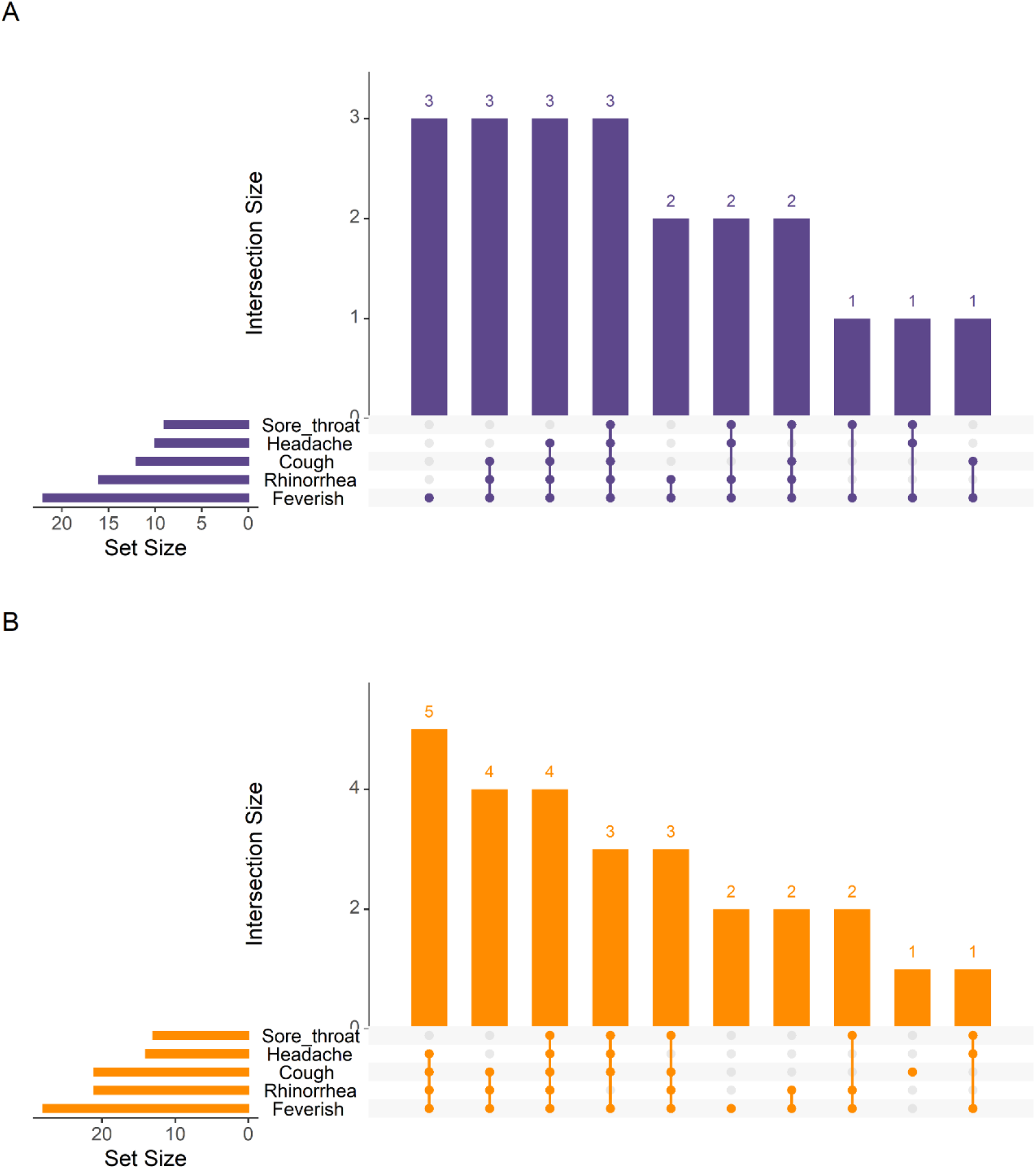
Comparison of Common Symptom Groupings between Symptomatic SARS-CoV-2 Infections in 2020 and 2021. A: 2020. B. 2021

## Discussion

This is, to our knowledge, the first study that assesses differences in symptom presentation, duration, and severity between SARS-CoV-2 and endemic HCoV symptomatic infections among children Understanding how SARS-CoV-2 infections compare to endemic HCoV infections is important as SARS-CoV-2 becomes endemic. Other studies have evaluated symptom presentation for endemic HCoV and SARS-CoV-2 infections in children separately. (2, 16–20) This work, however, compares medically attended illnesses associated with endemic HCoV and SARS-CoV-2 infections in a large, prospective cohort of children with high infection rates.

In this pediatric cohort, we found that with 854 (49%) participants tested positive for SARS-CoV-2 antibodies in 2021 with only 121 PCR confirmed infections that met the original testing criteria. These results are consistent with results from our community-based household cohort study in the same setting. (12) We also found that most participants had at least one alpha and one beta endemic HCoV infection by age four suggesting that in this cohort, participants have had at least two endemic HCoV exposures by the age of four. This is similar a previous study that showed that by age six, most children have had infections with each of the four endemic HCoVs with a majority being asymptomatic infections. (21) Thus for SARS-CoV-2 and endemic HCoV, symptomatic infections also represent only a small proportion of all pediatric infections.

Comparing these symptomatic infections, we found that pediatric disease presentation is very similar between endemic HCoVs and SARS-CoV-2, with each frequently presenting with “common cold” symptoms. Consistent with other studies, we found differences in symptom presentation by age; this may be, perhaps, because older children have had more endemic HCoV exposures (2, 4, 16, 17, 22). We also found great variability in symptom duration for SARS-CoV-2 and endemic HCoV infections; symptoms from endemic HCoV and SARS-CoV-2 infections lasted anywhere from 1 day to more than 28. (20) There was a difference in duration of loss of appetite for the youngest participants and cough for those aged 5-14 years suggesting that some symptoms may last longer for SARS-CoV-2 infections. We also found a difference in rhinorrhea duration between SARS-CoV-2 infections in 2020 and 2021, suggesting that SARS-CoV-2 variants may increase symptom duration among children.

Across age groups, risk of ALRI associated with SARS-CoV-2 was the same or lower compared to ALRI associated with endemic HCoV infection. Even when excluding endemic HCoV co-infections with pathogens commonly associated with increased risk of ALRI, the conclusions did not change. (16, 18) These results show that, for children, the risk of ALRI and severe illness from SARS-CoV-2 infections is comparable to the risk from endemic HCoV infections at the community level.

The main strength of this community-based study is its size and duration. Consistent viral surveillance and symptom evaluation within the same population allow for year-to-year comparisons and facilitates our comparisons of endemic HCoVs and SARS-CoV-2. The high number of participants under the age of five (about 36% of participants during these years), allows us to evaluate SARS-CoV-2 in an age group with little representation in current literature. Additionally, this cohort was already well established when SARS-CoV-2 began circulating in Nicaragua allowing us to quickly incorporate questions regarding its effects on this population.

However, this study does have some limitations. First, using data from this community-based cohort study we were not powered to detect the most severe manifestations of SARS-CoV-2 including death or Multisystem Inflammatory Syndrome in Children (MIS-C) and other rare outcomes. (23, 24) Second, our analysis did not include genetic sequencing preventing us from assessing the importance of variants in presentation and severity of SARS-CoV-2 illness. We did compare SARS-CoV-2 symptom presentation, severity, and duration by year and found little or no difference. Additionally, due to the low levels of circulation at the time, SARS-CoV-2 infections were only evaluated for influenza co-infections. We expect that RSV, and HMpV coinfections would also be associated with increased risk of ALRI for SARS-CoV-2; excluding such SARS-CoV-2 coinfections from the ALRI risk comparison would not change our findings. Finally, while our study does not evaluate very mild or asymptomatic illness for endemic HCoVs, our results were consistent with other research, showing that childhood HCoVs are ubiquitous and that symptomatic cases represent only a small proportion of infections, as with SARS-CoV-2. (21)

In this study, we observed that symptomatic SARS-CoV-2 infections at the community level are very similar to symptomatic endemic HCoV infections in symptom presentation. Among children in a tropical, urban setting with a high SARS-CoV-2 infection rate, symptomatic SARS-CoV-2 infections are on average as or less severe as endemic HCoV infections. These findings support the hypothesis that SARS-CoV-2 may be like another endemic HCoV for children—most children will be asymptomatic with rare cases of severe symptomatic illness. This does not mean SARS-CoV-2 in children is not important. There are many unknowns about the long-term effects and impact of repeat infections among children. (1) Increased transmissibility of emerging variants is also a cause for concern, as it will lead to increased frequency of severe manifestations. Future mutations in the virus needed to be monitored as they may also increase illness severity in children. Despite relatively low risk of severe illness among children, pediatric vaccination that mirrors natural induced immunity against SARS-CoV-2 would further lower individual risk and reduce the number of severe cases and deaths due to SARS-CoV-2.

## Data Availability

Data may be shared with outside investigators following UM IRB approval and once appropriate institutional approvals are obtained.

## Supporting information

**S1 Fig.**
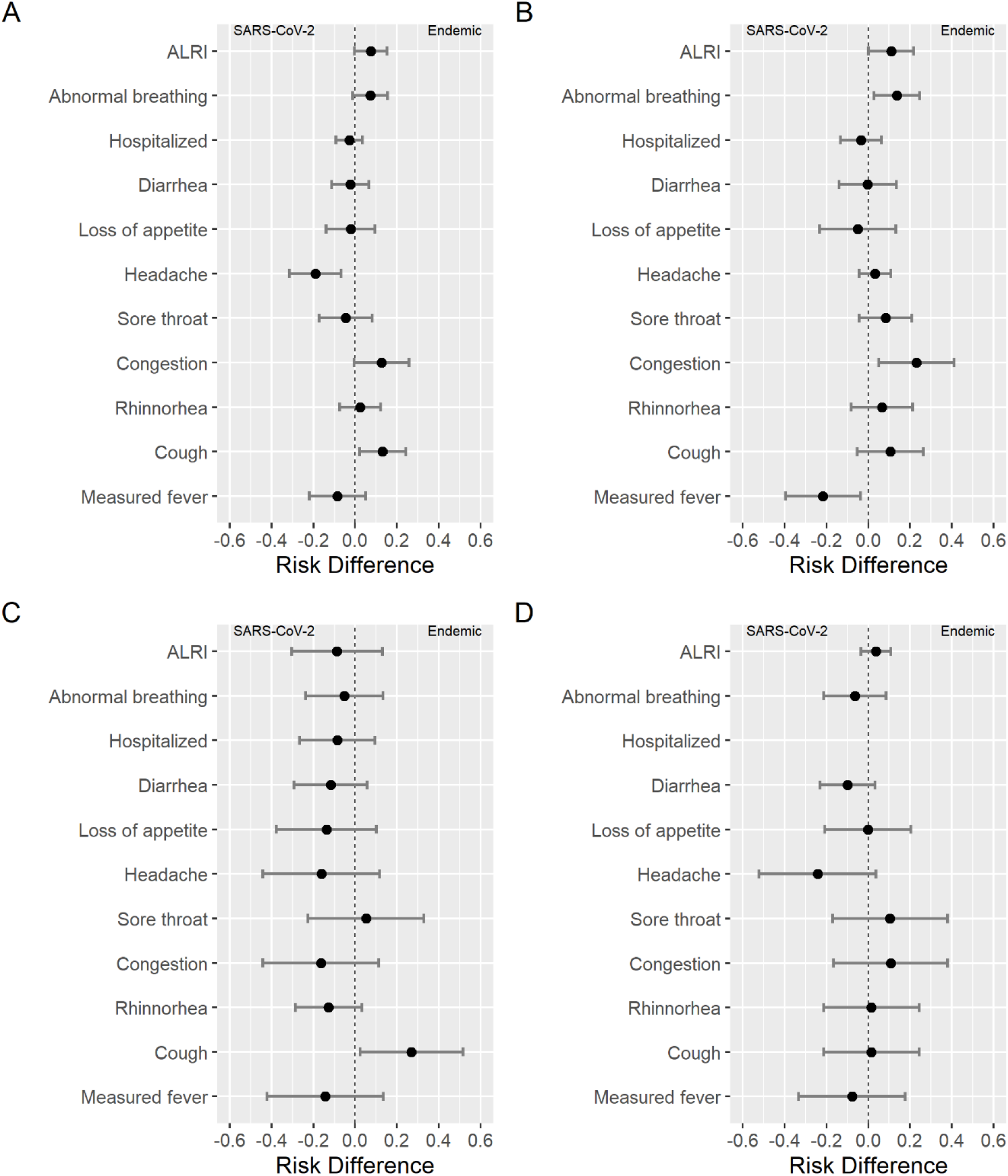
Symptom Risk Difference between Endemic HCoVs and SARS-CoV-2 for Females. A: All participants. B: Ages 0-4. C: Ages 5-9. D Ages: 10-14.

**S2 Fig.**
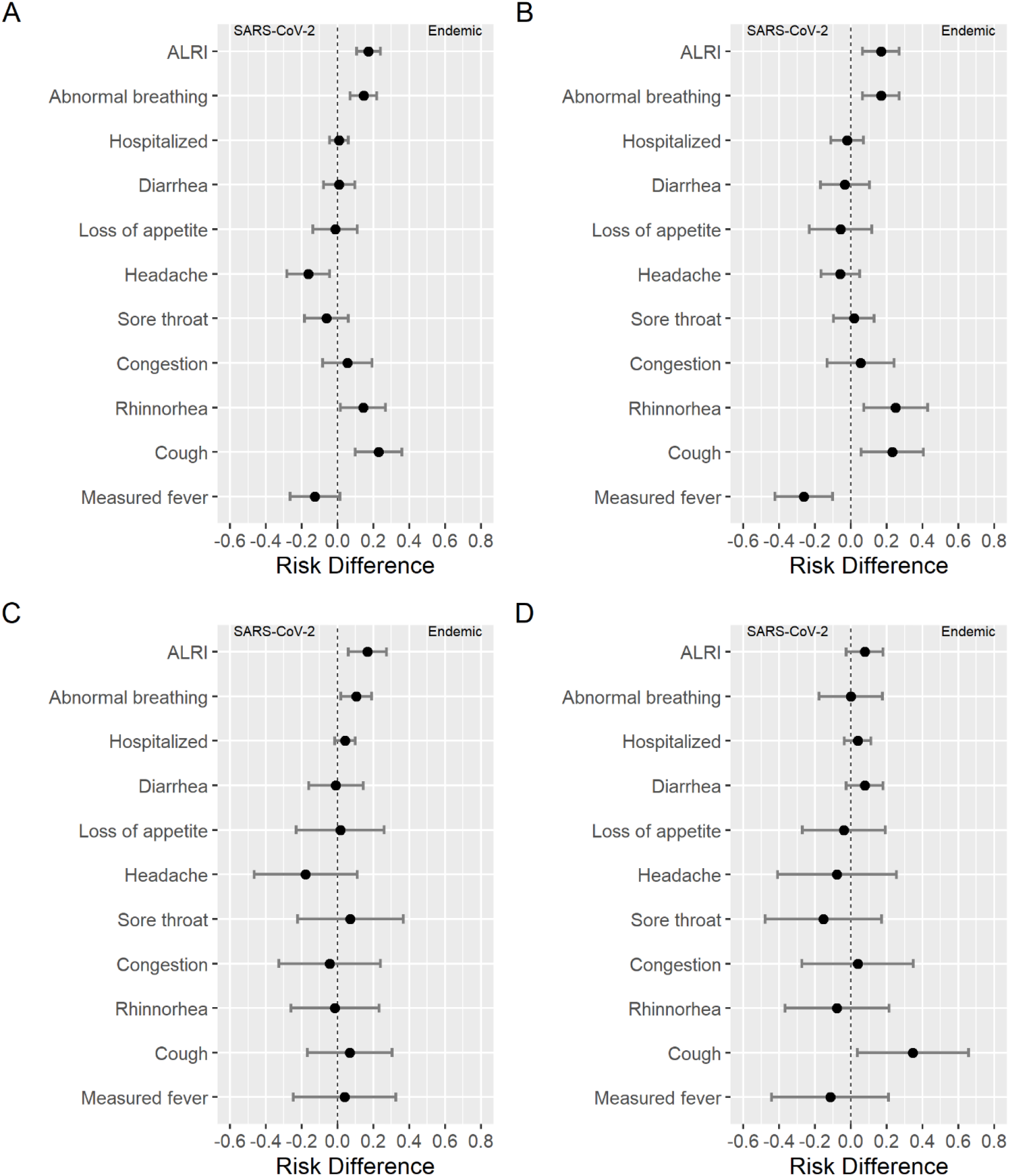
Symptom Risk Difference between Endemic HCoVs and SARS-CoV-2 for Males. A: All participants. B: Ages 0-4. C: Ages 5-9. D Ages: 10-14.

**S3 Fig.**
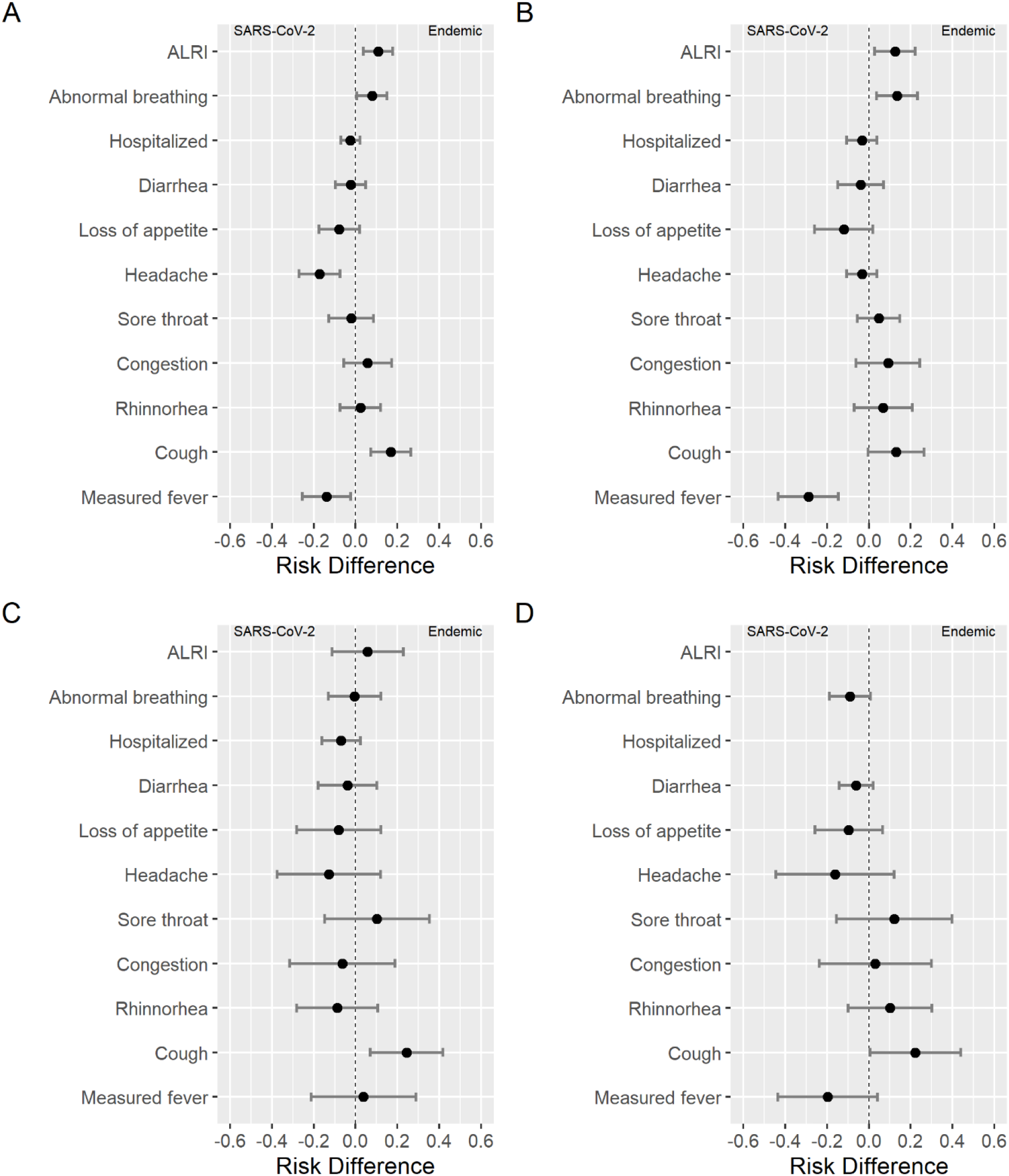
Symptom Risk Difference NL63 and SARS-CoV-2. A: All participants. B: Ages 0-4. C: Ages 5-9. D Ages: 10-14.

**S4 Fig.**
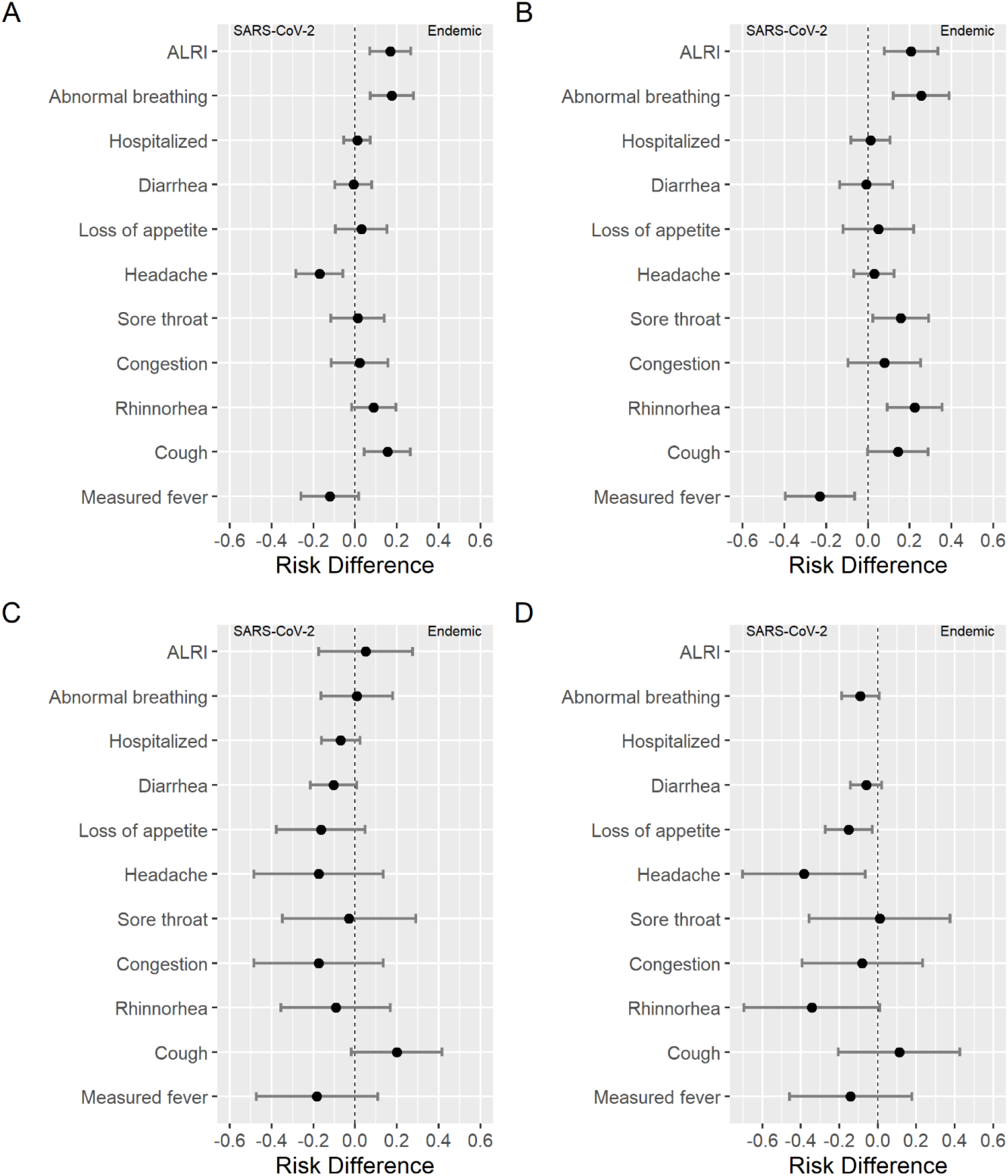
Symptom Risk Difference between 229E and SARS-CoV-2. A: All participants. B: Ages 0-4. C: Ages 5-9. D Ages: 10-14.

**S5 Fig.**
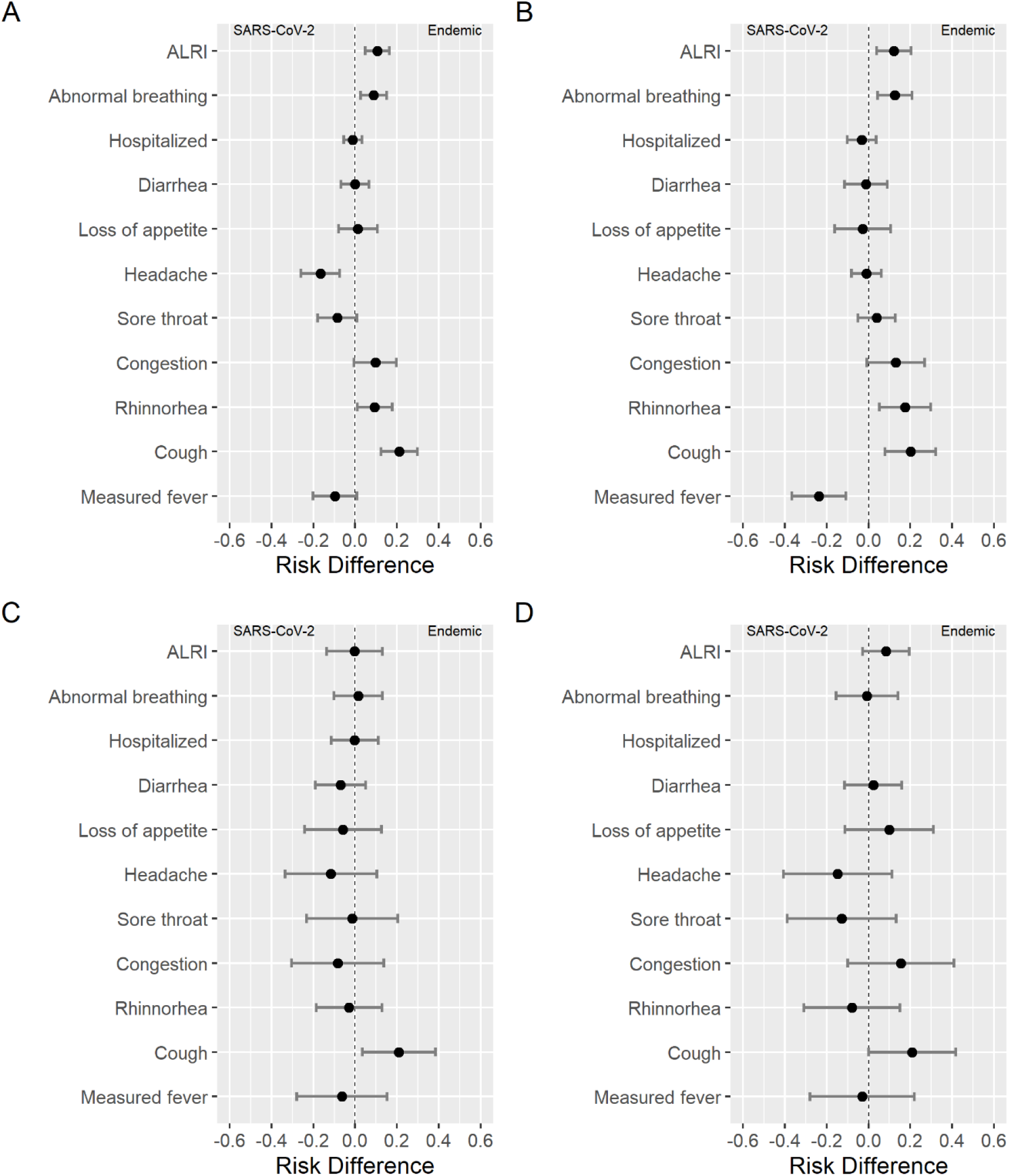
Symptom Risk Difference between OC43 and SARS-CoV-2. A: All participants. B: Ages 0-4. C: Ages 5-9. D Ages: 10-14.

**S6 Fig.**
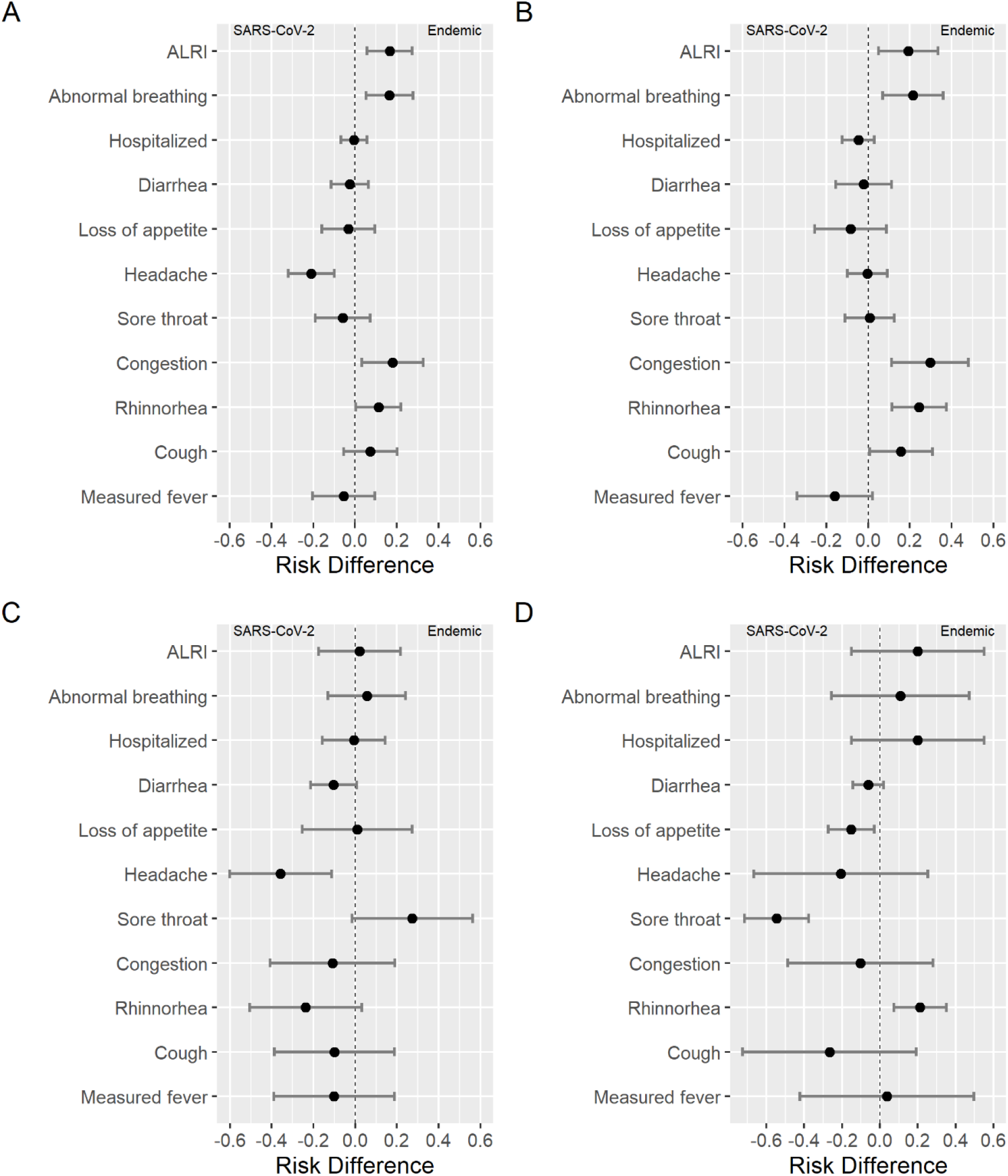
Symptom Risk Difference between HKU1 and SARS-CoV-2. A: All participants. B: Ages 0-4. C: Ages 5-9. D Ages: 10-14.

